# Improving Gonorrhoea Molecular Diagnostics: Genome Mining-Based Identification of Identical Multi-Repeat Sequences (IMRS) in *Neisseria gonorrhoeae* Genome

**DOI:** 10.1101/2023.07.20.23292966

**Authors:** Clement Shiluli, Shwetha Kamath, Bernard N. Kanoi, Rachael Kimani, Michael Maina, Harrison Waweru, Moses Kamita, Ibrahim Ndirangu, Hussein M. Abkallo, Bernard Oduor, Nicole Pamme, Joshua Dupaty, Catherine M. Klapperich, Srinivasa Raju Lolabattu, Jesse Gitaka

## Abstract

**Purpose:** Curable sexually transmitted infections (STIs) such as *Neisseria gonorrhoeae* (*N. gonorrhoeae*) is a major cause of poor pregnancy outcome. The infection is often asymptomatic in pregnant women and a syndrome-based approach of testing leads to missed diagnosis. Culture followed by microscopy is inadequate and time-consuming. The gold standard Nucleic Acid Amplification Tests (NAATs) require advanced infrastructure settings whilst point of care tests are limited to immunoassays with sensitivities and specificities insufficient to accurately diagnose asymptomatic cases. This necessitates the development and validation of assays that are fit for purpose.

**Materials and methods:** Here, we have identified new diagnostic target biomarker regions for *N. gonorrhoeae* using an algorithm for genome mining of identical multi repeat sequences (IMRS). These were then developed as DNA amplification primers to design better diagnostic assays. To test the primer pair, genomic DNA was 10-fold serially diluted (100pg/μL to 1×10^-3^pg/μL) and used as DNA template for PCR reactions. The gold standard PCR using 16S rRNA primers was also run as a comparative test, and both assay products resolved on 1% agarose gel.

**Results:** Our newly developed *N. gonorrhoeae* IMRS-PCR assay had an analytical sensitivity of 6 fg/μL representing better sensitivity compared to the 16S rRNA PCR assay with analytical sensitivity of 4.3096 pg/μL. The assay was also successfully validated with clinical urethral swab samples. We further advanced this technique by developing an iso-thermal IMRS, which was both reliable and sensitive for detecting cultured *N. gonorrhoeae* isolates at a concentration of 38 ng/μL. Combining the iso-thermal IMRS with a low-cost Lateral Flow Assay, we were able to detect *N. gonorrhoeae* amplicons at a starting concentration of 100 pg/μL.

**Conclusion:** Therefore, there is a potential to implement this concept within miniaturized, isothermal, microfluidic platforms, and laboratory-on-a-chip diagnostic devices for highly reliable point-of-care testing.

## 1 INTRODUCTION

Gonorrhoea, caused by *Neisseria gonorrhoeae* (*N. gonorrhoeae*) accounted for an estimated 82.4 million cases in 2020 globally [1]. Problems associated with *N. gonorrhoeae* infections include ectopic pregnancy, pelvic inflammatory disease and infertility [2].

Microscopy provides an affordable diagnostic option in symptomatic infections, but has low sensitivity in asymptomatic individuals [3] [4]. In men, *N. gonorrhoeae* microscopy is 84%–95% sensitive compared to culture and it is sensitive in symptomatic (90%–95%) than in asymptomatic (50%–75%) patients [5]. In female patients microscopic observation of cervical smears identifies approximately 50% of *N. gonorrhoeae* infections and is therefore not recommended for routine testing [6], [7] [8].

Antigen-based tests detect *N. gonorrhoeae* using antibodies immobilized on nitrocellulose strips [9]. However, data on the performance characteristics of these tests is limited [9].

Polymerase Chain Reaction (PCR) are sensitive but need expensive equipment, electricity, and cold chains for reagents [10][9] [11–13]. The high sensitivity of nucleic acid amplification technologies (NATs) such as the 16S rRNA PCR with a detection limit of 10 fg/μl in *N. gonorrhoeae* positive clinical samples is attributed to their ability to produce a positive test result from few copies of DNA or RNA. A real-time amplification assay based on the *N. gonorrhoeae opa* gene had a detection limit of 0.4 bacterial DNA copies [9]. This assay detects asymptomatic *N. gonorrhoeae* infections. However, the performance of this assay is yet to be validated on all of the *Neisseria* species [9]. Despite their many advantages, implementation of NAATs by laboratories is expensive and physical design constraints is a major challenge [9]. Also, NAATs tests such as the COBAS AMPLICOR test for example, produces false-positive results with non-pathogenic *Neisseria* species [14].

Sensitivity of PCR for detection of *N. gonorrhoeae* varies with different specimens. In a previous study, the sensitivity of PCR for gonorrhoea was 88.3% for urine specimens from men and 94.8% for urethral swab specimens from men [15]. A PCR based assay specific for the *orf*1 gene reported a high sensitivity of 25 fg DNA in clinical samples which is equivalent to 10 gonococcal cells [14].

Use of Loop-Mediated Isothermal Amplification (LAMP) test especially in low and middle-income countries has not been achieved. This is due to affordability and challenges with field deployment including need for cold chain storage of temperature sensitive reagents [10] [16] [17].

In this research, we developed a novel method based on *de novo* genome mining strategy that identifies repeat sequences in *N. gonorrhoeae* genome for use as both PCR and isothermal amplification coupled with lateral flow readout.

## 2 MATERIALS AND METHODS

### 2.1 IMRS genome mining algorithm

Primers were designed based on Identical Multi-Repeat Sequence (IMRS) genome mining algorithm as previously described [18]. The *Neisseria gonorrhoeae* (*N. gonorrhoeae*) genome (NZ_CP012028.1) was used as the reference template to identify identical, repetitive sequence substrings of <30 bases that can be used as forward or reverse primers. NIH’s Basic Local Alignment Search Tool (BLAST) was used to evaluate the primer pairs to confirm *N. gonorrhoeae* specificity.

### 2.2 Neisseria gonorrhoea Genomic DNA

*Neisseria gonorrhoea (N. gonorrhoeae*) DNA was obtained from the American Type Culture Collection (ATCC) (strain FA1090) (ATCC^®^ 700825DQ Batch No. 70035980) at a concentration of ≥1 x 10^5^ copies/μL. The stock DNA was diluted to a concentration of 100 pg/μL (3.5 x 10^4^ copies/μL) and thereafter serially diluted 100-fold and 10-fold in Tris-EDTA (TE) buffer (Thermo Fisher Scientific, Waltham, Massachusetts, USA) for the *N. gonorrhoeae* IMRS and 16S rRNA PCR, respectively.

### 2.3 16S rRNA PCR for *N. gonorrhoeae*

The 16S rRNA PCR assays were carried out as previously described [19]. Briefly, a reaction mixture containing dNTPs, (Thermo Fisher Scientific, Waltham, Massachusetts, USA) (0.2mM), forward and reverse primers (0.01 mM for each), Taq Hot-Start DNA polymerase (Thermo Fisher Scientific, Waltham, Massachusetts, USA) (1.25 U), genomic template DNA, 1 μL to a final PCR reaction of 25 μL was used. The cycling parameters for *N. gonorrhoeae* 16S rRNA PCR were as follows: 95 °C for 3 min; 30 cycles of: 95 °C for 30 s, 53 °C for 30 s, 72 °C for 30 s; and 72 °C for 5 min and a final hold of 4 °C. Non-template controls were included in all the PCR assays and were also included in the gel electrophoresis.

### 2.4 N. gonorrhoeae IMRS PCR

The *N. gonorrhoeae* identical multi-repeat sequence (*N. gonorrhoeae*-IMRS) assays were carried out as previously described [18] in a reaction mixture containing dNTPs, (Thermo Fisher Scientific, Massachusetts, USA) (0.2 mM), forward and reverse primers (0.01 mM for each), Taq Hot-Start DNA polymerase (Thermo Fisher Scientific, Massachusetts, USA) (1.25 U), genomic template DNA, 1 μL to a final PCR reaction of 25 μL. The cycling parameters for *N. gonorrhoeae*-IMRS assay was as follows: 95 °C for 3 min; 30 cycles of: 95 °C for 30 s, 55 °C for 30 s, 72 °C for 30 s; and 72 °C for 5 min and a final hold of 4 °C.

All PCR products were resolved on a 1% agarose gel visualized on a UV gel illuminator system under ethidium bromide staining. Non-template controls were included in all the PCR assays and were also included in the gel electrophoresis.

### 2.5 Isothermal IMRS Amplification Assay

The Isothermal (Iso) IMRS amplification were performed in a 25 μL reaction mixture as previously described [20] and consisted of *Bst* 2.0 polymerase (640 U/mL) (New England Biolabs, Massachusetts, USA), with 1× isothermal amplification buffer, 3.2 μM forward primer, and 1.6 μM reverse primer (Jigsaw Biosolutions, Bengaluru, India) combined with 10 mM dNTPs (Thermo Fisher Scientific, Massachusetts, USA), 0.4 M Betaine (Sigma-Aldrich, Missouri, USA), molecular-grade water and Ficoll (0.4 g/mL) (Sigma-Aldrich, Missouri, USA). Amplification was carried out at 56°C for 40 min. Amplified products were visualized by gel electrophoresis in a 1% gel. Non-template controls were included in all the PCR assays and were also included in the gel electrophoresis.

### 2.6 Lower limit of detection

To assess the lower limit of detection (LLOD), for the *N. gonorrhoeae* IMRS PCR assay, genomic DNA was diluted 100 folds from 10^2^ pg/μL (3.5 x 10^4^ copies/μL) to 10^-6^ pg/μL (<1 copies/ μL) and 10 folds from 10^2^ pg/μL (3.5 x 10^4^ copies/μL) to 10^-2^ pg/μL (3.5 copies/ μL) for the gold standard 16S rRNA PCR. Thereafter, five replicates of each dilution were used for the assays. Amplification products were visualized on a 1% gel. To determine the LLOD of the *N. gonorrhoea* 16S rRNA and IMRS PCR, probit analysis was performed using the ratio of successful reactions to the total number of reactions performed for each assay.

### 2.7. Culture of *Neisseria gonorrhoea (N. gonorrhoeae)* isolates and DNA extraction

Urethral *N. gonorrhoeae* positive isolates, confirmed by microscopy, were inoculated on Modified Thayer Martin (MTM) agar plates (Thermo Fisher Scientific, Kansas, USA) and streaked for isolation. The plates were incubated at 36°C in 5% CO_2_. Culture plates were examined for colonies typical of *N. gonorrhoeae* daily for up to 72 h. Cultures were confirmed as *N. gonorrhoeae* by characteristic gonococcal colonial morphology and the oxidase biochemical tests. DNA extraction was done using the Qiagen kit (Hilden, Germany) and thereafter, quantification was done using the Nanodrop spectrophotometer instrument.

### 2.8 Cloning and characterization of *N. gonorrhoeae*-IMRS amplicons

Gene cloning was performed as previously described [20] to confirm the sequences of amplicons obtained from the *N. gonorrhoeae*-IMRS PCR assay. *Neisseria gonorrhoea (*N. gonorrhoeae) gDNA was amplified using Assembly_IMRS-F (ttccggatggctcgagtttttcagcaagat<u>GTTGCCCCGCCCCGGCTCAAAGG)</u> and Assembly_IMRS-R (agaatattgtaggagatcttctagaaagat<u>ATAGCGGATTAACAAAAATCAGGACAAGGC</u>) primers. The underlined uppercase sequences correspond to the IMRS primers for amplifying the *N. gonorrhoeae* genome whereas the non-priming overlap lowercase sequence at the 5 - end of the primers sequence corresponds to the homologous sequences in the cloning vector. The resulting ∼223 bp amplicons were resolved on a 2% agarose gel to confirm the fragment size and subsequently purified using the PureLink™ PCR purification kit (ThermoFisher). The purified amplicon was then ligated into pJET1.2 blunt vector (ThermoFisher) using the NEBuilder® HiFi DNA Assembly kit (NEB) as per the manufacturer’s instruction. The resulting NEBuilder HiFi DNA Assembly product was transformed into NEB 5αCompetent *E. coli* (#C2987, NEB) following the manufacturer’s instructions. Transformed colonies were randomly selected, DNA extracted and Sanger-sequenced using the universal pJET1.2 forward sequencing primer (CGACTCACTATAGGGAGAGCGGC) and pJET1.2 forward sequencing primer (AAGAACATCGATTTTCCATGGCAG). The resulting nucleotides were trimmed and analyzed using SnapGene software (GSL Biotech; available at snapgene.com), aligned to check for similarity or “clonal” differences and BLAST was used to check for similarity with the *N. gonorrhoeae* genome.

### 2.9 *N. gonorrhoeae*-Lateral Flow Assay

To generate a visual read-out signal of amplicons, an *N. gonorrhoeae*-Lateral Flow Assay (*N. gonorrhoeae*-LFA) was used as previously described [20]. Briefly, 2.5 μL annealing buffer, dNTPs and NaCl 1.75 μL, MgSO_4_ 1.2 μL *N. gonorrhoeae* 5’ biotinylated forward primer, *N. gonorrhoeae* Reverse primer, *N. gonorrhoeae* 3’ FAM labelled probe, N. gonorrhoeae Digotexin labelled probe, Ficoll 400 6.25 μL, *N. gonorrhoeae* internal control sequence DNA 2.5 μL, ISO Amp III enzyme mix 2.0 μL, template DNA 5 μL and molecular grade water 1.45 μL in a final master mix volume of 25 μL was used. Incubation of the LFA strip (Milenia Biotec GmbH, Giessen, Germany) was performed at 65°C for 1 h and 5 μL of reaction mixture was then added to the LFA strip, thereafter, 2 drops of running buffer was also added.

### 2.10 Test for *N. gonorrhoeae* IMRS PCR primers interference with human DNA

Approximately 3 mL venous blood was collected from three donors and used to prepare dried blood spots (DBS) on Whatman filter paper. The DBS were incubated for 24 h at room temperature. The DBS were cut into 6-mm diameter circles using a single-hole punch, and DNA was extracted using the QIAamp DNA mini extraction spin column, yielding a final DNA volume of 100 μL, following the manufacturer’s instructions (Qiagen, Hilden, Germany). Extracted DNA was then used as template for the *N. gonorrhoeae*-IMRS PCR assay using previously mentioned amplification conditions. The *N. gonorrhoeae* genomic DNA was used as a positive control. Non-template controls were included in all the PCR assays and were also included in the gel electrophoresis.

### 2.11 *N. gonorrhoeae* IMRS and 16S rRNA PCR real-time PCR assay

The Quant Studio 5 Real-Time PCR System (with Quant Studio Design and Analysis Desktop Software v1.5.) was used as a reference method [21] for the *N. gonorrhoeae*-16S rRNA PCR and the *N. gonorrhoeae*-IMRS-PCR and for determining the sensitivity of the *N. gonorrhoeae* IMRS and 16S rRNA PCR primers for detecting *N. gonorrhoeae*. Serially diluted (starting concentration of 10^3^ genome copies/μL serially diluted 100 folds for *N. gonorrhoeae*-IMRS and starting concentration of 10^4^ genome copies/μL serially diluted 10-folds for 16S rRNA) genomic DNA were used as template. The final real-time PCR master mix volume was 10 μL in triplicate and consisted of the following components; 5 μL SYBR Green qPCR Master Mix (Thermo fisher, Massachusetts, USA), 1 μL forward and reverse IMRS primer mix, 2.5 μL template genomic DNA and 1.5 μL molecular grade water. The amplification cycling conditions were 50°C for 2 min; 95°C for 10 min; 40 cycles of 95°C for 15 s and 60°C for 30 s.

### 2.12 Clinical samples and ethics statement

Vaginal samples from women enrolled in an STI screening study at the Kenyatta National Hospital in Nairobi County were used in this study. The *N. gonorrhoeae*-16S rRNA conventional PCR assay was used to confirm *N. gonorrhoeae* infections.

### 2.12 Data analysis

Graphs were plotted with GraphPad Prism version 7.0 (GraphPad Software, San Diego, CA). The mean, and Standard Deviation values were calculated with Excel 2016. To determine the LLOD of *N. gonorrhoeae*-IMRS and *N. gonorrhoeae*-16S rRNA PCR assays (the concentration at which genomic *Neisseria gonorrhoea* DNA is detected with 95% confidence), probit regression analyses were performed in Excel 2016. Genomic *Chlamydia trachomatis, Trichomonas vaginalis* and *Treponema pallidum* DNA were used to determine the specificity of the *N. gonorrhoeae*-IMRS primers for other STIs. *P* < 0.05 was considered as significant.

## 3 RESULTS

### 3.1 Design, and distribution of IMRS primer sets on *Neisseria gonorrhoea* genome

A total of 16 repeats (Table 1) were identified using the IMRS-based genome mining algorithm, which could be used as forward and reverse primers for an amplification assay. These selected primer pair (*Ng*IMRS-Primer-F 5’- GTTGCCCCGCCCCGGCTCAAAGG- 3’, *Ng*IMRS-Primer-R 5’- ATAGCGGATTAACAAAAATCAGGACAAGGC - 3’) were identified on the genome sequence of *Neisseria gonorrhoeae* 35/02 strain (GenBank accession number CP012028, version: CP012028). As depicted in the Circos plot in Figure 1A, the primers were found to be present at various loci of the sense and antisense strands, allowing them to serve interchangeably as forward or reverse primers.

**Figure 1:**
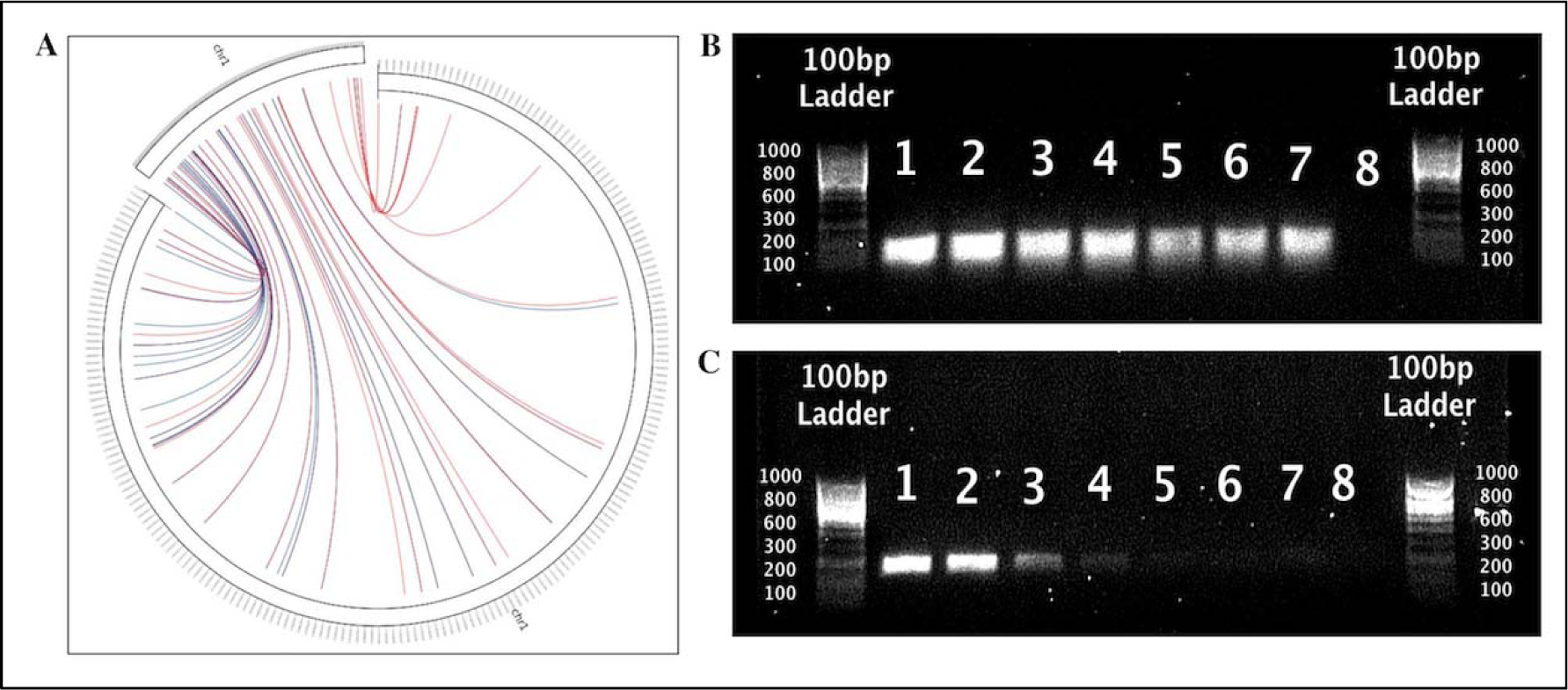
*N. gonorrhoeae*-IMRS Primer targets on the genome and gel image of the *N. gonorrhoeae*-IMRS and 16S rRNA PCR assay. (A) Circos plot for the distribution of identical multi repeat sequence (IMRS) primers in the *Neisseria gonorrhoeae* genome. *Neisseria gonorrhoeae* IMRS primer A (blue lines) and *Neisseria gonorrhoeae* IMRS primer B (red lines) both have 16 repeats. Image of 10-fold serially diluted (1 – 100, 2 – 10, 3 – 1, 4 – 0.1, 5 – 0.01, 6 – 0.01, 7 – 0.001 and 8 - NTC) (pg/μL) genomic *Neisseria gonorrhoeae* DNA amplicons resolved on a 1% gel using IMRS primers (B) and gold standard 16S rRNA PCR (C). The final length of PCR products obtained using IMRS primers was 124 bp and gold standard 16S rRNA PCR was 260 bp.

**Table 1:** *N. gonorrhoeae*-IMRS primer targets on the bacterial genome.

| *N. gonorrhoeae*-IMRS Primer amplification regions |  |
| --- | --- |
| 1 | 277972 - 278095 |
| 2 | 442143 - 442266 |
| 3 | 534149 - 534272 |
| 4 | 659675 - 659798 |
| 5 | 741065 - 741188 |
| 6 | 1170515 - 1170638 |
| 7 | 1350817 - 1350941 |
| 8 | 1818578 - 1818701 |
| 9 | 2064281 - 2064404 |
| 10 | 505504 - 505381 (-) |
| 11 | 825946 - 825823 (-) |
| 12 | 924241 - 924118 (-) |
| 13 | 960240 - 960117 (-) |
| 14 | 978398 - 978275 (-) |
| 15 | 1858932 - 1858809 (-) |
| 16 | 1971573 - 1971450 (-) |

### 3.2 Amplification of genomic *Neisseria gonorrhoea* using identical multi repeat and gold standard 16S rRNA PCR primers

The primers were used to amplify serially diluted *N. gonorrhoeae* DNA genome. The IMRS primers could detect *N. gonorrhoeae* genomic DNA from a concentration of as low as 1 fg/μL (Fig 1B), whereas the gold standard 16S rRNA primers (Fig 1C) could only detect *N. gonorrhoeae* genomic DNA down to a concentration of 1 pg/μL. This demonstrates that the IMRS-based PCR assay has a higher sensitivity compared to the gold standard 16S rRNA PCR assay. Real-time PCR was also performed using serially diluted genomic *N. gonorrhoeae* DNA as a template and *N. gonorrhoeae*-IMRS primers as well as conventional *N. gonorrhoeae*-16S rRNA primers. The mean Ct values at each respective dilution were used to plot graphs (Fig 2C for *N. gonorrhoeae*-16S rRNA primers and for *N. gonorrhoeae*-IMRS primers).

**Figure 2:**
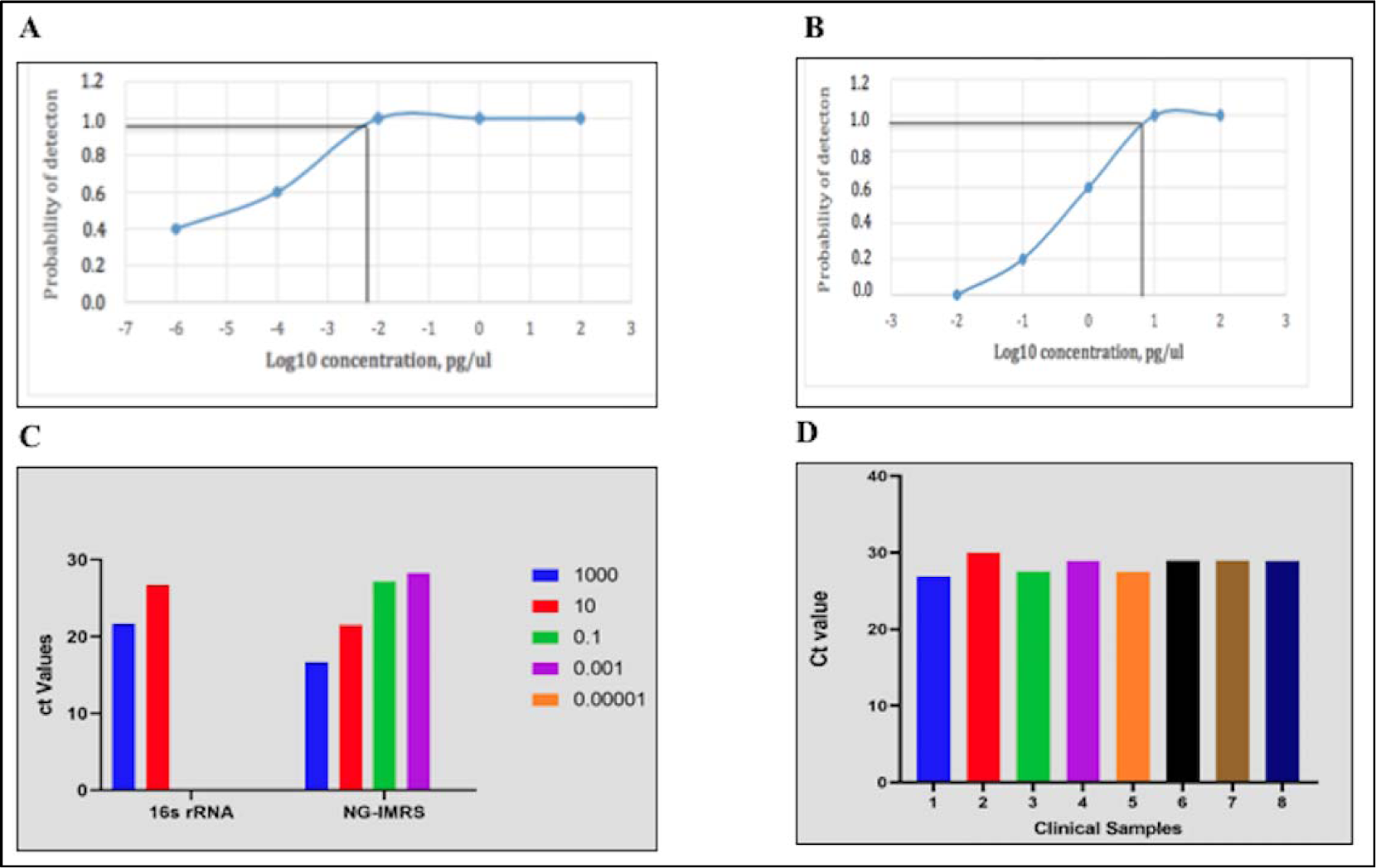
Probit analysis estimation for IMRS. As indicated (A), the IMRS primers for *Neisseria gonorrhoeae* had an LLOD = 6 fg/μL. B: Probit analysis estimation for 16S rRNA PCR. As indicated, gold standard primers for *Neisseria gonorrhoeae* had an LLOD = 4.3096 pg/μL. C: Plots showing Ct values obtained from amplification of genomic *Neisseria gonorrhoeae* serially diluted genomic DNA using RT-PCR for the 16S rRNA and *N. gonorrhoeae*-IMRS primers. The legend shows the starting concentration of the genomic DNA dilutions. D: A total of eight PCR confirmed *Neisseria gonorrhoeae* clinical samples were used to validate the *N. gonorrhoeae*-IMRS primers using RT-PCR.

### 3.3 Calculation of the lower limit of detection (LLOD) for IMRS PCR and gold standard 16S RNA PCR

To determine the lowest limit of detection (LLOD) of the IMRS PCR relative to the gold standard 16S rRNA PCR, probit statistic was performed using *N. gonorrhoeae* genomic DNA serially diluted 100-fold (Table 2A) and 10-fold (Table 2B) and used as template for the *N. gonorrhoeae*-IMRS and 16S rRNA PCR respectively. Fig 2A shows the probit plot for the *N. gonorrhoeae*-IMRS PCR assay, and Fig 2B shows the probit plot for the gold standard 16S rRNA PCR assay. The LLOD was calculated as the concentration at which *N. gonorrhoeae* DNA can be detected with 95% confidence. Probit analysis estimation for *N. gonorrhoeae*-IMRS PCR, Coefficient χ = −5.0456, *P*-value 0.0095 (Table 2C). As indicated, the IMRS primers for *Neisseria gonorrhoea* had an LLOD = 6 fg/μL, Fig 3A. Probit analysis estimation for 16S rRNA PCR χ = −7.1656, *P*-value 0.9976, Table 2C. As indicated, gold standard primers for *Neisseria gonorrhoea* had an LLOD = 4.31 pg/μL, Fig 2B. Table 2C shows the statistics obtained from the Probit analysis indicating that the *N. gonorrhoeae*-IMRS PCR assay had increased sensitivity compared to the gold standard 16S rRNA PCR assay.

**Figure 3:**
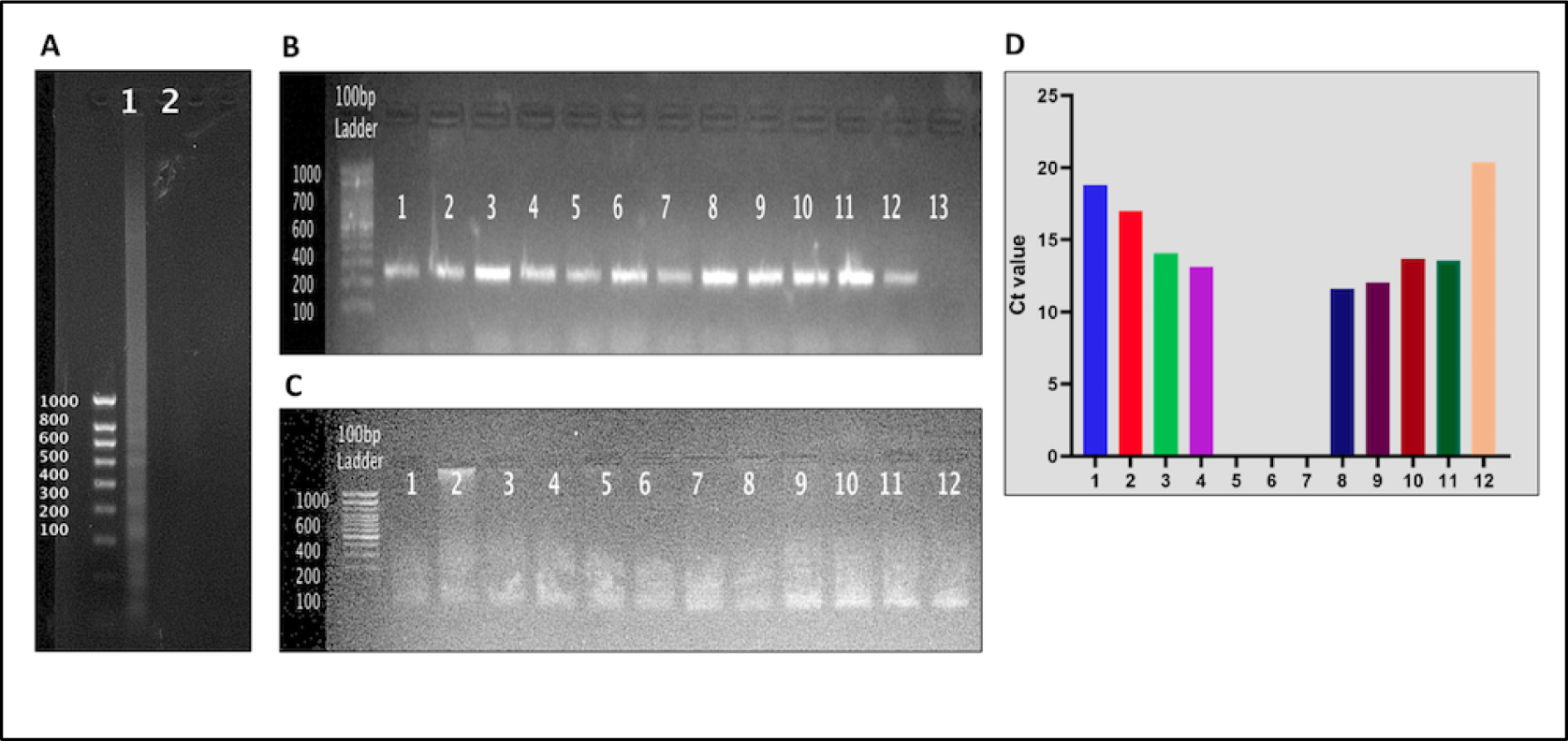
Gel images of DNA amplified using the Isothermal *N. gonorrhoeae*-IMRS primers (A) and the 16S rRNA (B) and the *N. gonorrhoeae* IMRS PCR (C) and Ct values (D) obtained from the RT-PCR assay using cultured *Neisseria gonorrhoeae* clinical isolates *N. gonorrhoeae*-IMRS primers were used in the RT-PCR. Amplification products were resolved on a 1% gel. I – sample, 2, Negative control, (Figure A). 1 – 12 samples, 13 – Negative control (Figure B and C). The *N. gonorrhoeae*-IMRS assay had a sensitivity of between 66 – 100% and a specificity of 71%.

**Table 2:**
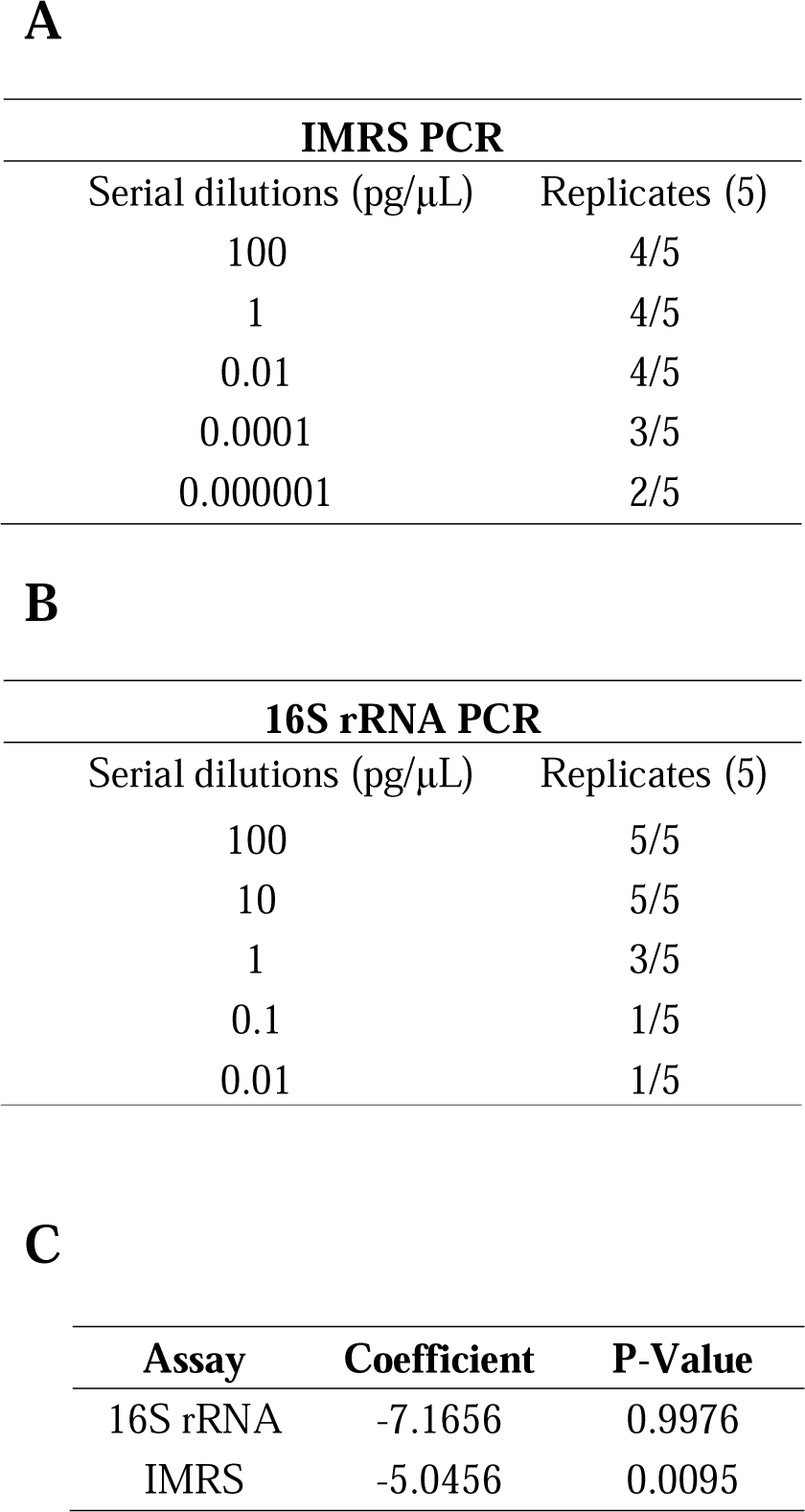
*Neisseria gonorrhoeae* genomic DNA was serially diluted 100 folds (A) and 10-folds (B) and used as template for the *N. gonorrhoeae*-IMRS and 16S rRNA PCR to estimate the lower limit of detection. Table (C) shows the statistics obtained from the Probit analysis.

### 3.4 Isothermal amplification of cultured *Neisseria gonorrhoea* DNA

Extracted DNA (38 ng/μL) from cultured *Neisseria gonorrhoea* was used to perform ISO-IMRS. As shown in Figure 3A, the reaction products were visualized on a 1% gel.

### 3.5 Validation of *N. gonorrhoeae* IMRS PCR assays with clinical samples

The *N. gonorrhoeae* IMRS primers were validated with DNA isolated from 12 culture confirmed clinical samples. The 16S rRNA PCR assay was run as a control and amplicons resolved on a 1% gel, Fig 3B. As shown in Fig 3C, the *N. gonorrhoeae* IMRS PCR assay successfully amplified DNA extracted from the 12 culture confirmed samples tested. Using real-time PCR as a reference, three of the cultured samples were scored as false positive results (Fig 3D). A total of eight *N. gonorrhoeae* positive DNA samples were used to evaluate the reliability of the *N. gonorrhoeae*-IMRS primers for detecting *N. gonorrhoeae* DNA using RT-PCR assay as shown in Fig 2D. The *N. gonorrhoeae*-IMRS primers showed concordance with the results obtained from PCR diagnosis, successfully identifying *N. gonorrhoeae* infections in infected samples.

### 3.6 Cloning and sequencing of *N. gonorrhoeae*-IMRS amplicons

DNA from transformed *E. coli* cells was extracted and sequenced (Fig 4C). Multiple sequence alignment confirmed that the sequences matched *N. gonorrhoeae* deposited on publicly available resources at *in silico* predicted genomic coordinates. These results confirmed that the *N. gonorrhoeae* IMRS primers were specific for targets within the *N. gonorrhoeae* genome. In addition, we designed a probe specific to amplified sequences within the *N. gonorrhoeae* genome. As indicated in Fig 4D, the *N. gonorrhoeae*-IMRS probe was specific to serially diluted *N. gonorrhoeae* genomic DNA.

**Figure 4:**
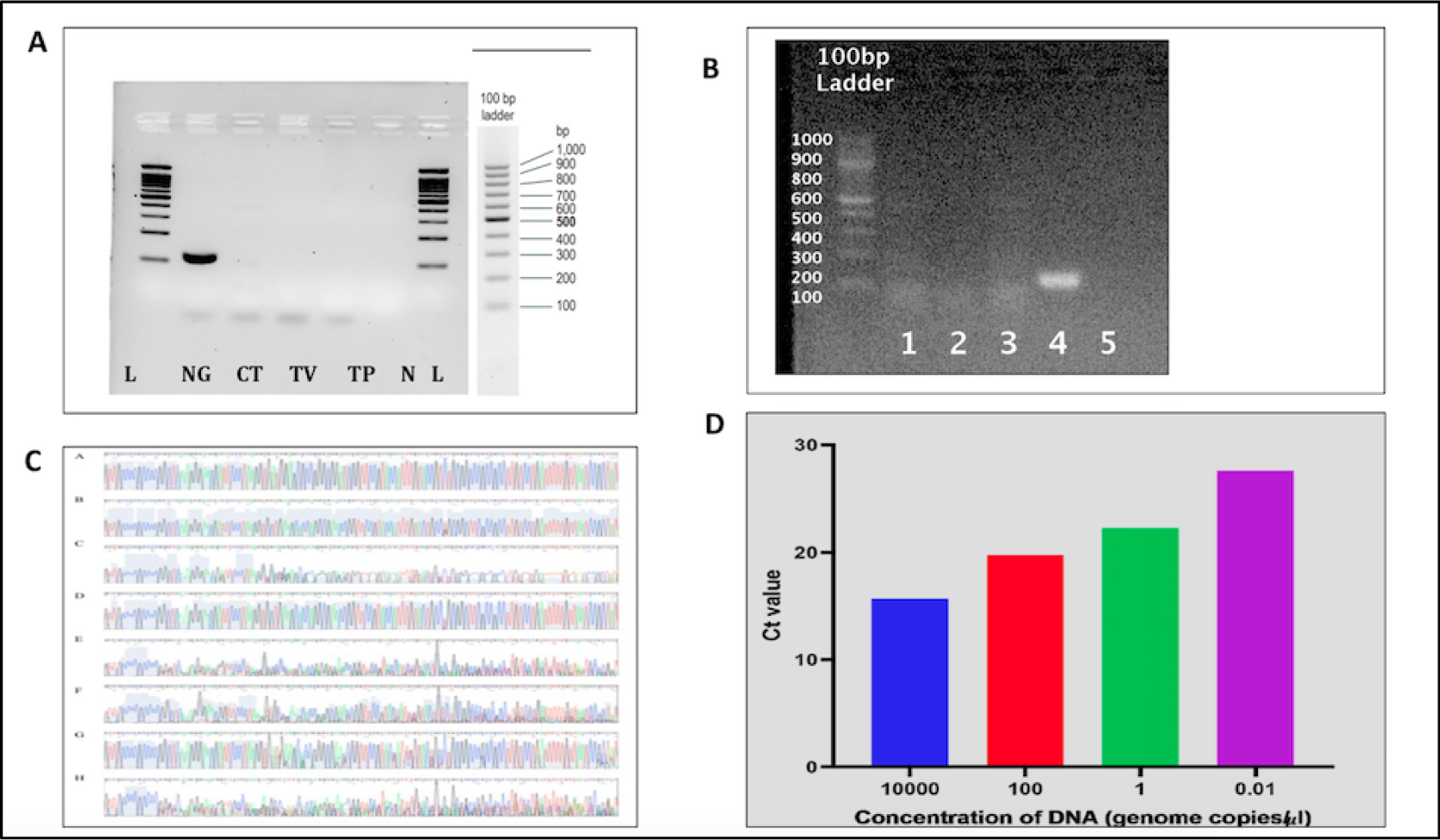
To determine the specificity of *N. gonorrhoeae*-IMRS primers, *Chlamydia trachomatis, Treponema pallidum* and *Trichomonas vaginalis* genomic DNA was used as PCR template (A). (B) Gel image of human DNA extracted from venous blood collected from donors (1 - 3). The extracted DNA served as template for the *N. gonorrhoeae*-IMRS PCR assay and PCR products were resolved on a 1% gel as shown. 6, Positive control – genomic *N. gonorrhoeae* DNA, 7 – Negative control. (C) *Neisseria gonorrhoeae* sequences (A-124bp, B-125bp, C-118bp, D-124bp, E-123bp, F-123bp, G-124bp and H-122bp). (D) Ct values from RT-PCR using a probe specific to *Neisseria gonorrhoeae* sequences.

### 3.7 Specificity of *N. gonorrhoeae*-IMRS primers and interference with Human DNA

As shown in Fig 4A, the *N. gonorrhoeae*-IMRS primers were specific only for *Neisseria gonorrhoea* DNA and non-specific for *Treponema pallidum* (TP), *Chlamydia trachomatis* (CT) and *Trichomonas vaginalis* (TV) genomic DNA. *N. gonorrhoeae* genomic DNA was used as a positive control. The *N. gonorrhoeae*-IMRS primers were also non-specific to *Neisseria meningitides* (Supplementary data). Also, compared to the gold standard conventional 16S rRNA PCR, the *N. gonorrhoeae*-IMRS PCR reliably detected genomic DNA at a concentration of 1 fg/μL (Fig 1B). The *N. gonorrhoeae*-IMRS primers were also tested for interference with DNA extracted from human whole blood. As shown in Fig 4B, the *N. gonorrhoeae*-IMRS primers were non-specific to human DNA.

### 3.8 *Neisseria gonorrhoea* Lateral Flow Assay (LFA)

A visual readout of diluted genomic DNA was developed using modified primers and labelled probes, as shown in Fig 5. The LFA readout of the amplification products was successful indicating the potential for the Iso-IMRS-based assay to be used in field settings.

**Figure 5:**
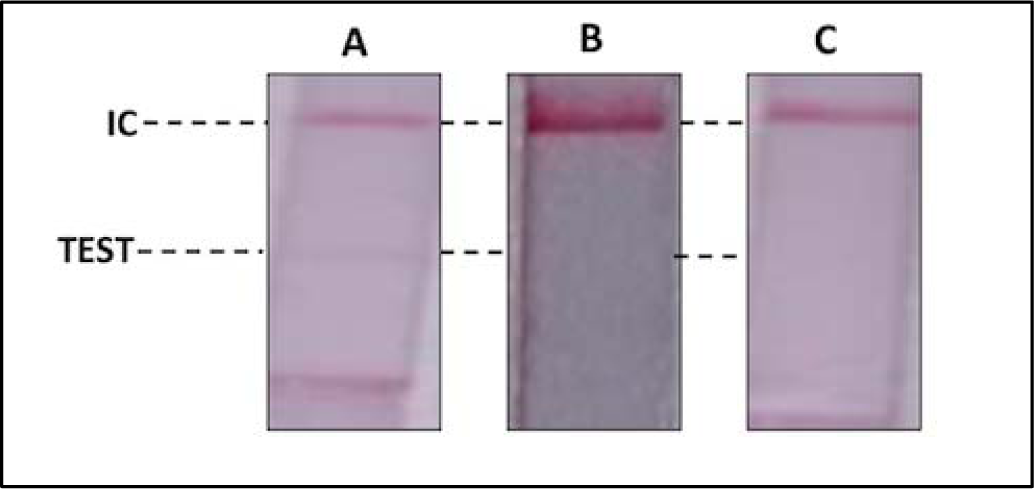
Detection of serially diluted *Neisseria gonorrhoeae* DNA. (A): 100pg/μl, (B):10pg/μL, (C): Non Template Control. Strips were incubated at 65°C for 1 h IC – Internal Control.

## 4 DISCUSSION

A key priority in the control of STIs is the development of point-of-care tests that help overcome the existing barriers of diagnosis [22]. Compared to the 16S rRNA PCR, amplification of specific sequences on *N. gonorrhoeae* genome using IMRS primers was more sensitive generating a large number of amplicons of varying sizes from the genomic DNA as shown in Fig 1B. The IMRS PCR assay was more sensitive with a lower limit of detection of 6 fg/μL (2.5 copies/μL, Fig 2A) representing approximately 1052 times better sensitivity. A previous study based on the Loop-Mediated Isothermal Amplification (LAMP) of Meningococcal genomic DNA reported a lower limit of 10 genome copies per reaction [23]. Compared to the LAMP assay, the *N. gonorrhoeae*-IMRS assay yielded a higher detection limit

Using real-time PCR to determine the sensitivity of the *N. gonorrhoeae*-IMRS PCR method, we were able to detect up to 1 genome copies/mL of *N. gonorrhoeae* genomic DNA. Efficient detection of asymptomatic *N. gonorrhoeae* infection has been identified as a priority in high burden countries. In this perspective, it would be necessary to evaluate the sensitivity of the *N. gonorrhoeae*-IMRS assay in large number of clinical samples.

A study that used real-time PCR assay to target the gonococcal 23S rRNA C2611T and A2059G mutations from vaginal swabs reported Ct value ranges of 29.87 – 37.98 and 29.2 – 39.44 respectively [24]. Using clinical samples to validate the *N. gonorrhoeae*-IMRS assay, we obtained Ct value ranges of between 27 and 29 (Fig 2D), indicating increased sensitivity of the novel assay.

We further showed that isothermal amplification using IMRS PCR primers detected cultured *N. gonorrhoeae* DNA. The concentration of the template was 38ng/μL (Fig 3). Our findings are similar to a study that showed a novel Isothermal-IMRS method was specific for detection of gonococcal strains carrying *penA*-60.001 ceftriaxone resistant allele [25].

Using *N. gonorrhoeae* genomic DNA as a positive control cross-species specificity for the *N. gonorrhoeae*-IMRS primers was determined using *Chlamydia trachomatis, Trichomonas vaginalis* and *Treponema pallidum* genomic DNA (Fig 4A). Cloning of amplicons followed by sequencing with the plasmid-specific primers yielded the reads that aligned within the expected *N. gonorrhoeae* genomic sequences thus confirming the amplicons were generated as expected (Fig 4C).

In the present study, we have also developed a LFA for the diagnostic testing of *N. gonorrhoeae* in low resource settings. Our *N. gonorrhoeae*-LFA can detect amplicons at a concentration of 100pg/μl (Fig 5) visible to the naked eye. The *N. gonorrhoeae*-LFA was more sensitive than a gold nanoparticles colorimetric readout test that detected 10^6^ CFU/mL in whole live cells in culture [26]. The *N. gonorrhoeae*-LFA can therefore replace syndromic management or culture-based clinical methods for the diagnosis of Gonorrhoea*e* particularly in low-resource settings.

## 5 CONCLUSION

The designed assay can potentially be used to screen asymptomatic infections due to its increased sensitivity and specificity. Our ongoing studies with other STI pathogens indicate that the IMRS algorithm-based genome mining can yield biomarkers for STIs.

## Data Availability

All data produced in the present work are contained in the manuscript

## CRediT author statement

C.S.: conceptualization, methodology, formal analysis, writing—original draft; S.K.: data curation, formal analysis, investigation, methodology, supervision, visualization, writing—original draft, writing—review and editing; B.N.K.: investigation, writing—review and editing; R.K.: resources, writing—review and editing; M.M.: resources, writing—review and editing; H.W.: resources, writing—review and editing; M.K.: resources, writing—review and editing; I.N.: resources, writing—review and editing; H.M.A.: resources, writing— review and editing; B.O.: funding acquisition, writing—review and editing; N.P.: resources, writing—review and editing;. J.D.: writing—review and editing; C.M.K.: resources, writing—review and editing; S.R.L.: methodology, writing—review and editing; J.G.: conceptualization, funding acquisition, investigation, methodology, project administration, resources, supervision, writing—original draft, writing—review and editing.

## Ethical Statement

Use of these samples and study proposal was approved on the 23^rd^ of October 2020 by the Mount Kenya University Ethical Review Committee (MKU/ERC/1649).

## Acknowledgement

We thank Dr. Karen Muthembwa, Mr. Meshack Juma and all clinicians working at the STI Clinic in Kenyatta National Hospital.

## Data Accessibility

All data generated or analysed during this study are included in this publishedarticle [and its supplementary information files].

---

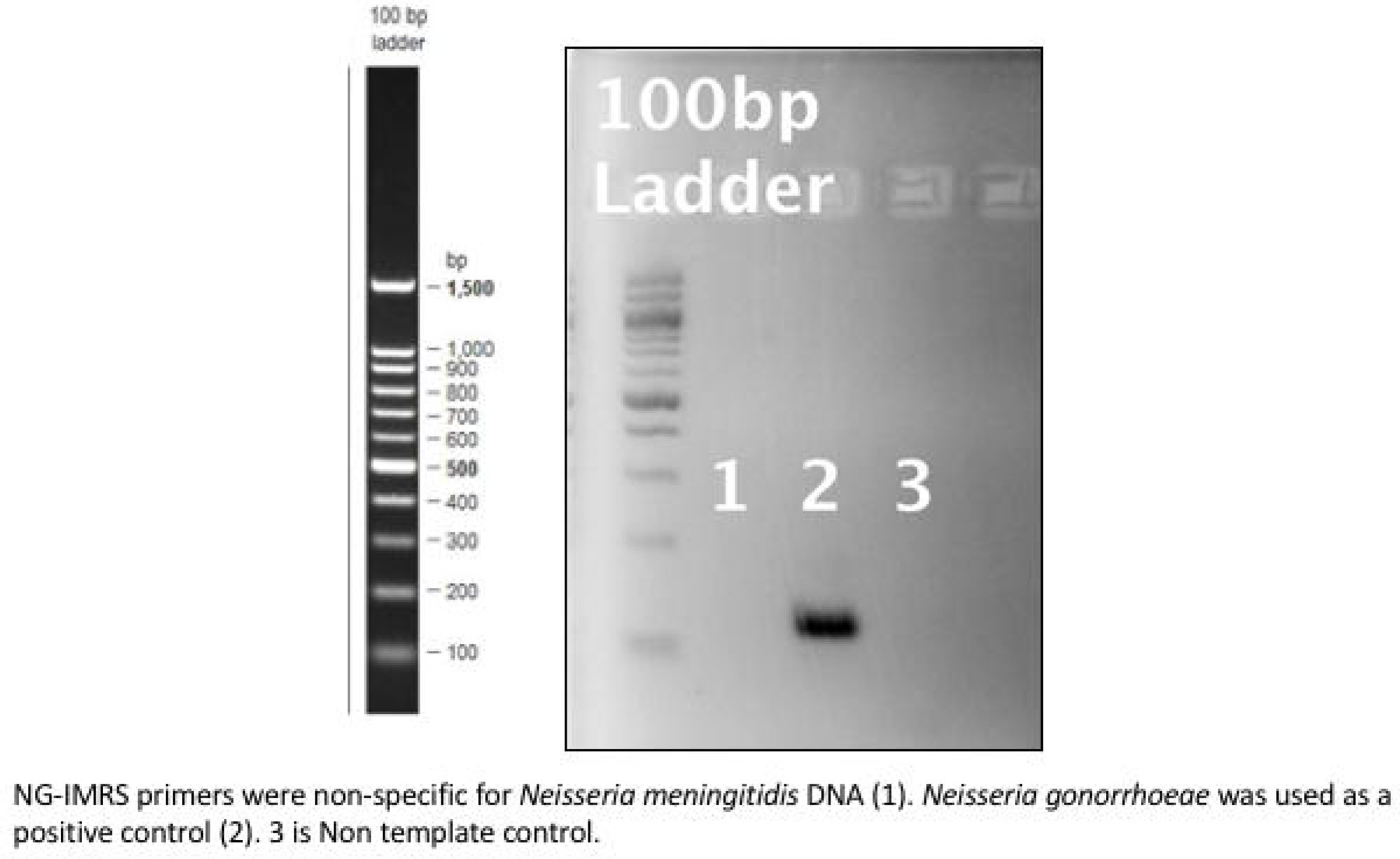

